# COVID-19 Vaccine Acceptability and Inequity in the United States: Results from a nationally representative survey

**DOI:** 10.1101/2021.01.29.21250784

**Authors:** Dustin Gibson, Smisha Agarwal, Ankita Meghani, Rupali J. Limaye, Alain Labrique

**Author notes:** Corresponding author’s address Dustin Gibson, Johns Hopkins Bloomberg School of Public Health 615 N Wolfe St, Rm E8650, Baltimore, MD 21205.

## Abstract

**Background:** At the time of this survey, September 1^st^, there were roughly 6 million COVID-19 cases and 176,771 deaths in the United States and no federally approved vaccine. The objective of this study was to explore the willingness to accept a COVID-19 vaccine in the United States and describe variability in this acceptability by key racial, ethnic and socio-demographic characteristics.

**Methods:** This was a cross-sectional digital survey that sampled participants from a nationally-representative panel maintained by a third party, Dynata. Dynata randomly sampled their database and emailed web-based surveys to United States residents ensuring the sample was matched to US Census estimates for age, race, gender, income, and Census region. Participants were asked how willing or unwilling they would be to: 1) receive a COVID-19 vaccine as soon as it was made publicly available, and 2) receive the influenza vaccine for the upcoming influenza season. Participants could respond with extremely willing, willing, unwilling, or extremely unwilling. For those who reported being unwilling to receive a COVID-19 vaccine, reasons for this hesitancy were captured. All participants were asked about where they obtain vaccine-related information, and which sources they trust most. Univariable and multivariable logistic regressions were conducted to examine the association of all demographic characteristics with willingness to receive COVID-19 vaccine.

**Findings:** From September 1^st^ to September 7, 2020, 1592 respondents completed the online survey. Overall, weighted analyses found that only 58.9% of the sample population were either willing or extremely willing to receive a COVID-19 vaccine as soon as it was made publicly available. In comparison, 67.7% of the respondents were willing or extremely willing to take the influenza vaccine. By gender, 66.1% of males and 51.5% of females were willing to receive a COVID-19 vaccine. Males were significantly more willing to receive a COVID-19 vaccine (adjusted odds ratio (OR)=1.98, 95% CI: 1.56, 2.53; p<0.001) than females. Blacks were the least willing racial/ethnic group (48.8%) Blacks, (aOR=0.59, 95%CI: 0.43, 0.80; p<0.001) were significantly less willing, than whites, to receive a COVID-19 vaccine. There were numerous reasons provided for being unwilling to receive a COVID-19 vaccine. The most common reason was concern about the vaccine’s safety (36.9%), followed by concerns over its efficacy (19.1%).

**Interpretation:** In conclusion, we found that a substantial proportion (41%) of United States residents are unwilling to receive a COVID-19 vaccine as soon as one is made publicly available. We found that vaccine acceptance differs by sub-populations. In addition to sub-group differences in willingness to receive the vaccine, respondents provided a variety of reasons for being unwilling to receive the vaccine, driven by various sources of vaccine information (and misinformation). This compounds the challenge of delivering a safe and efficacious COVID-19 vaccine at a population level to achieve herd immunity. A multi-pronged and targeted communications and outreach effort is likely needed to achieve a high level of immunization coverage.

## Introduction

The coronavirus disease 2019 (COVID-19) pandemic has killed more than 1.5 million people and infected roughly 70 million people worldwide.^1^ According to Johns Hopkins University, the United States accounts for more than a fifth of these infections, the most cases of any nation. COVID-19 has also caused 283,658 deaths in the United States with more than 2,000 deaths being reported per day as of December 07, 2020. Since November 2020, a record number of new cases have been reported, saturating hospital beds and in many cases exceeding the intensive care unit capacity across the country.^2^

While non-pharmaceutical interventions (NPI), such as, social distancing measures, school closures, stay-at-home orders, quarantine and isolation, use of face masks, and other hygiene behaviors, and therapeutics have helped manage and control disease transmission, they have been insufficient for containing COVID-19.^3^ Many factors have contributed to the heterogenous application of NPIs across US states and over time, including political resistance and shifting federal guidance to states. National-level implementation of NPIs have also resulted in economic strife and disproportionately affected marginalized populations, especially among African American and ethnic minority populations across the country. Given the high estimated case fatality rate of 1.8%,^4^ an effective vaccine with sufficient population-level coverage is the safest way to achieve herd immunity again SAR-CoV-2. If sufficiently large portions of the population are vaccinated against COVID-19, then transmission slows, making it possible to protect unvaccinated populations, and individuals who cannot be vaccinated, e.g., immunocompromised persons or newborns.

Several pharmaceutical and biotechnology companies are developing potential vaccine candidates, supported by global and national initiatives to accelerate the discovery to production lifecycle. The urgent global need for an efficacious vaccine has spurred companies and public health agencies to compress vaccine development and review timelines, gaining speed through process efficiencies and massive financial investments. As of December 2020, seven vaccines have already been approved for early or limited use worldwide. These include the two-dose Pfizer-BioNTech vaccine, and the Moderna vaccine, that have shown a vaccine efficacy of 95% and 94.1% respectively.^5,6^ Both are expected to receive an emergency use authorization (EUA), paving the way for vaccine administration to start in December 2020.

Though several safe and effective vaccine candidates may receive FDA authorization by early 2021, achieving herd immunity requires a sufficiently large portion of the population to accept getting vaccinated. In the United States, experts estimate that 70% of the population would need to be vaccinated to halt COVID-19 transmission.^7^ Recent data suggest that COVID-19 vaccine acceptance is suboptimal, as a study in May reported that only 67% of those surveyed would accept the vaccine (ref). In the United States, efforts to contain the COVID-19 pandemic have been highly politicized, creating an erosion of public trust through waves of misinformation around the effectiveness of mask-wearing to the safety of COVID-19 vaccines. The accelerated pace from discovery to development of these vaccines have raised public concerns around whether shortcuts might have compromised the safety of the vaccine candidates.^8^

Historically, in the US, there has been increasing reluctance to accepting new vaccines,^9^ with significant variations in attitudes towards immunization by race and ethnicity.^10–12^ Willingness to accept a COVID-19 vaccine also varies by socio-demographic characteristics, with increased acceptance by males, older adults, those who identify as Asian Americans, and those with higher education.^8^ Empirical data indicate stark racial disparities with regards to willingness to accept vaccines.^10–12^ In the case of the seasonal influenza vaccine, Blacks and Hispanics have lower rates of coverage compared to Whites. There are also marked disparities of vaccine coverage by age^13^ as higher coverage is observed among older adults (>65 years and older) relative to younger adults.^14^ In addition to financial and socioeconomic challenges to access healthcare in these often marginalized populations, a history of formal medical exploitation and abuse of these communities of color continues to provoke mistrust and fear – a consequence of decades of institutional and cultural racism.^15^

The objective of this study was to explore the willingness to accept a COVID-19 vaccine in the United States and describe variability in this acceptability by key racial, ethnic and socio-demographic characteristics. Our goal is to use findings from this study to provide policymakers and public health practitioners with relevant and timely information to develop targeted messaging to facilitate COVID-19 vaccine uptake and reduce disparities in coverage. Given the disproportionate burden of death and morbidity in these populations, this research is critical to the rapid and effective control of the US COVID-19 pandemic.

## Methods

### Study setting

Survey participants were recruited nationally, across the US, from September 1st to September 7^th^, 2020. By late August, the COVID-19 pandemic in the US had reached a new phase as cases were becoming more widespread as compared to the concentrated outbreaks that had been observed earlier in the pandemic.^16^ By September 1^st^, nationwide there were roughly 6 million COVID-19 cases (confirmed and probable),^17^ and 176,771 deaths (confirmed and probable).^18^ Data revealed a disproportionately higher burden of COVID-19 cases and deaths among African American, Hispanic, and American Indian and Alaska Natives,^19^ casting a spotlight on long-standing health system and access inequities within the US population. Rural communities suffered from shortages in clinical personnel, protective gear, and even intensive care unit beds.^20^ Also in the early Summer months, the leading COVID-19 vaccine manufacturers began the recruitment for Phase 3 clinical trials,^21^ with the United States federal government securing advance purchase agreements with large vaccine manufacturers if their vaccines proved efficacious and were approved by regulatory agencies.^22^ The study protocol and survey instruments were approved by the Institutional Review Board at Johns Hopkins Bloomberg School of Public Health (IRB00012413). All participants provided consent (electronically).

### Study Design

This was a cross-sectional digital survey that sampled participants from a nationally-representative panel maintained by Dynata (https://www.dynata.com). Dynata’s database of more than 62 million unique users with accompanying demographic information has been previously used to conduct state- and nationally-representative COVID-19 surveys.^10^ Dynata employs a series of quality control measures include digital fingerprinting to prevent duplication, spot checking via third party verification to prove identity, reward redemption quality procedures, and benchmarking against known external data points.

### Procedures

Given a targeted set of socio-demographic characteristics, Dynata randomly sampled their database and emailed web-based surveys to United States residents ensuring the sample was matched to US Census estimates for age, race, gender, income, and Census region. Participants were ineligible for the survey if they self-reported an age less than 18 years old or were not currently living in the United States. Dynata provided a small monetary remuneration to participants upon survey completion.

The survey consisted of questions on: 1) demographics, 2) risk perceptions, 3) education, 4) viral testing, 5) stigma and agency, 6) COVID-19 knowledge, 7) medication and treatment perceptions, 8) pregnancy and antenatal care and 9) vaccine perceptions and information. For the vaccine module, participants were asked how willing or unwilling they would be to: 1) receive a COVID-19 vaccine as soon as it was made publicly available, and 2) receive the influenza vaccine for the upcoming influenza season. Participants could respond with extremely willing, willing, unwilling, or extremely unwilling. For those who reported being unwilling to receive a COVID-19 vaccine, reasons for this hesitancy were captured. All participants were asked about where they obtain vaccine-related information, and which sources they trust most.

### Statistical Analyses

Sample size estimates calculated that 1004 participants were needed to produce population estimates with a margin of error of plus or minus three percentage points. We inflated the sample for Blacks and Hispanics, to 385 participants in each group to be able to detect a 10% difference in proportions between the racial groups (e.g., 40 to 50%).

Willingness to receive a COVID-19 vaccine and the influenza vaccine were collapsed into binary variables by combining willing and extremely willing responses and by combining unwilling and extremely unwilling choices. Overall percentages were presented, unweighted and weighted, and stratified by self-reported race (White, Black, or Hispanic), age group, income level, gender, census region, political party affiliation and education level. Weights for race by Census region were generated using 2010 Census data. Univariable and multivariable logistic regressions were conducted to examine the association of all demographic characteristics with willingness to receive COVID-19 vaccine. Participants who indicated either being unwilling or extremely unwilling were asked to provide a reason for why they would not be vaccinated. Pearson’s Chi-squared tests were used to examine differences in distribution of these reasons, and also sources of vaccine information, by socio-demographic characteristics. We used standard definitions for disposition codes and survey rates from the American Association of Polling and Opinion Research (AAPOR).^23^ Participants were classified as Non-Contacts if they either did not initiate the survey or failed to answer the age question. Partial interviews (PI) were defined as age-eligible participants who did not complete the survey. Complete interviews (I) were defined as people who finished the entire questionnaire. We used AAPOR equation #2 and #1 for response and cooperation rates, respectively.^23^

### Role of the funding source

The funder of the study had no role in study design, data collection, data analysis, data interpretation, or writing of the report. The corresponding author had full access to all the data in the study and had final responsibility for the decision to submit for publication.

## Results

From September 1^st^ to September 7, 2020, email invitations were sent to 16,904 potential participants. Participants under the age of 18 years (n=47), those not currently living in the United States (n=3), or those who provided a race whose required sample quota was filled (n=171) were ineligible for the survey. Seventy-two participants opened the survey but did not provide electronic consent; 77 participants were considered partial interviews. The survey’s response and cooperation rate were, respectively, 10.0% and 95.3%. The following are results from 1,592 complete interviews (I). Socio-demographic characteristics, unweighted and weighted for race by Census region are found in Table 1. White participants were more likely to have a graduate degree, be aged 35-54 years old, and affiliate with the Republican party than Blacks and Hispanics (See Appendix Table 1).

**Table 1.**
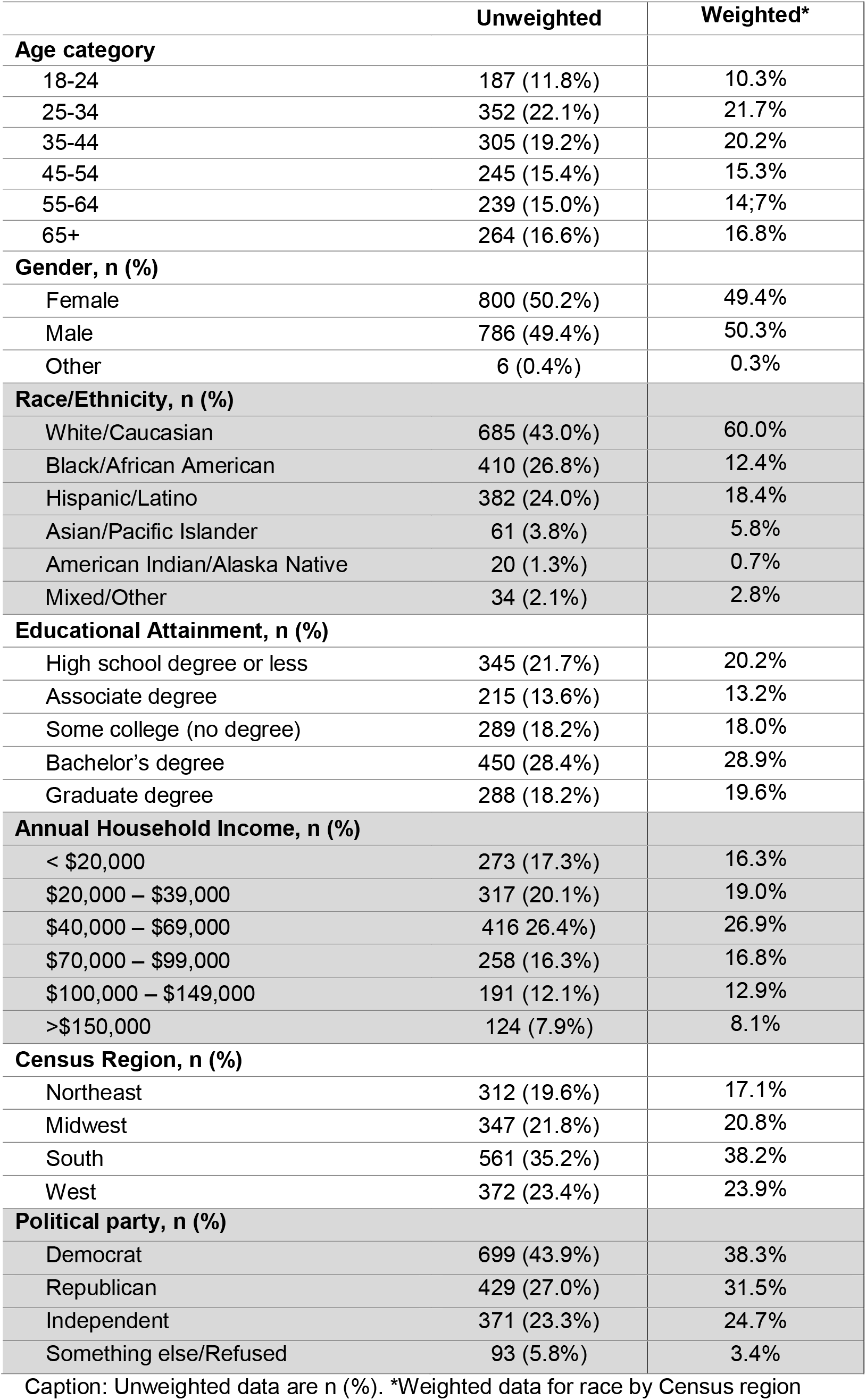
Demographic characteristics of complete interviews (n=1592)

Overall, weighted analyses found that only 58.9% of the sample population were either willing or extremely willing to receive a COVID-19 vaccine as soon as it was made publicly available. In comparison, 67.7% of the respondent group were willing or extremely willing to take the influenza vaccine. Sub-group analyses for weighted prevalence of willingness to receive COVID-19 vaccine are presented in Figure 1 and Appendix 2. By gender, 66.1% of males (95%CI: 62.4%, 69.7%) and 51.5% of females (95% CI: 47.6%, 55.4%) were willing to receive a COVID-19 vaccine. Participants aged 45-54 years (52.4% [95%CI: 45.4 - 59.2%]) and 55-64 years (50.4% [95% CI 43.4-57.5%]) were least willing to receive a COVID-19 vaccine. African Americans were the least willing racial/ethnic group (48.8%, 95%CI: 43.7%, 54.0%). By education, participants with a graduate degree had the highest levels of willingness (70.3%, 95%CI: 64.2%, 75.7%). Aside from the Midwest (53.6%, [95%CI 47.5% - 59.7%]), all Census regions had estimates of willing to accept the vaccine above the national average of 58.9%. Participants earning less than 20,000 dollars (51.8%, 95%CI: 44.9%, 58.6%) and independents (51.7%, 95%CI, 46.1%, 57.2%) were least willing to receive COVID-19 vaccine. Willingness to receive a COVID-19 vaccine was generally lower than willingness to receive an influenza vaccine across all sub-groups (See Appendix 3)

**Figure 1.**
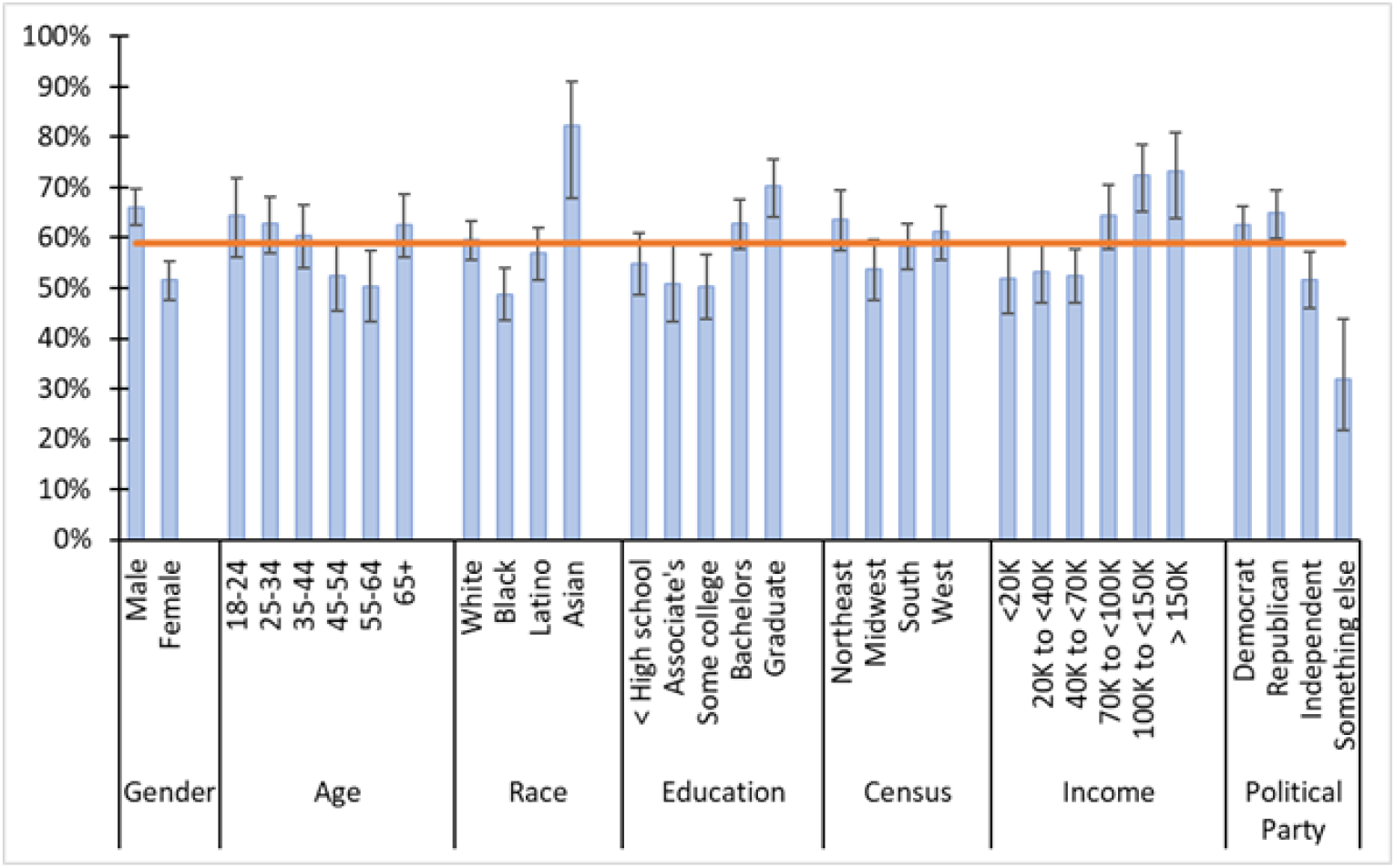
Willingness to receive COVID-19 vaccine when publicly available by subgroups. Red horizontal line is national average (58.9%) for willingness to receive a COVID-19 vaccine as soon as publicly available

In the multivariable analyses (Table 2), as compared to the youngest age group, 18-24 year-olds, participants aged 35-44 years (aOR=0.51, 95%CI 0.32,0.82, p=0.0060), 45-54 year-olds (aOR=0.43, 95% CI: 0.26, 0.70, p=0.0006), and 55-64 year-olds (aOR=0.42, 95%CI 0.26, 0.68, p=0.0005) were significantly less willing to receive a COVID-19 vaccine. Males were significantly more willing to receive a COVID-19 vaccine (aOR=1.98, 95% CI: 1.56, 2.53; p<0.001) than females. With regards to race, Blacks, (aOR=0.59, 95%CI: 0.43, 0.80; p<0.001) were significantly less willing, than whites, to receive a COVID-19 vaccine. Asians were more likely (aOR=3.19, 95%CI: 1.45, 7.01; p=0.004) than whites to receive a COVID-19 vaccine Participants in the highest income brackets, those earning $100,000 to $149,999 (aOR=1.70, 95%CI: 1.25, 2.31; p=0.0008) and more than $200,000 (aOR=1.70, 95%CI: 1.25, 2.31; p=0.0008) were significantly more likely than the lowest income group, households earning less than $20,000, to be willing to receive the COVID-19 vaccine. Those identifying as being politically independent were less willing (aOR=0.62, 95%CI: 0.45, 0.86; p=0.004) to receive a COVID-19 vaccine as compared to Republicans. There were no significant differences in willingness to receive a COVID-19 vaccine between Republicans and Democrats as well as no significant differences by education level and Census region in the multivariate analysis.

**Table 2.** Univariable and Multivariable logistic regression models for demographic characteristics associated with willingness to receive COVID-19 vaccine.

|  | OR (95% CI) | p-value | aOR (95% CI) | p-value |
| --- | --- | --- | --- | --- |
| **Age group (year)** |  |  |  |  |
| 18-24 | *Ref.* | . | *Ref.* | . |
| 25-34 | 0.93 (0.61, 1.41) | 0.729 | 0.66 (0.42, 1.03) | 0.067 |
| 35-44 | 0.85 (0.55, 1.30) | 0.441 | 0.51 (0.32, 0.82) | 0.006 |
| 45-54 | 0.61 (0.39, 0.95) | 0.029 | 0.43 (0.26, 0.70) | <0.001 |
| 55-64 | 0.56 (0.36, 0.88) | 0.012 | 0.42 (0.26, 0.68) | <0.001 |
| 65+ | 0.92 (0.60, 1.42) | 0.717 | 0.66 (0.41, 1.06) | 0.087 |
| **Gender** |  |  |  |  |
| Female | *Ref.* | . | *Ref.* | . |
| Male | 1.84 (1.46, 2.31) | <0.001 | 1.98 (1.56, 2.53) | <0.001 |
| **Race** |  |  |  |  |
| White | *Ref.* | . | *Ref.* | . |
| Black | 0.65 (0.50, 0.84) | 0.001 | 0.59 (0.43, 0.80) | <0.001 |
| Latino | 0.90 (0.69, 1.17) | 0.428 | 0.86 (0.64, 1.16) | 0.333 |
| Asian | 3.17 (1.41, 7.12) | 0.005 | 3.19 (1.45, 7.01) | 0.004 |
| American Indian | 1.18 (0.25, 5.67) | 0.836 | 1.00 (0.13, 7.56) | 0.999 |
| Other | 0.76 (0.36, 1.59) | 0.466 | 0.92 (0.42, 1.97) | 0.815 |
| **Education** |  |  |  |  |
| Highschool or less | *Ref.* | . | *Ref.* | . |
| Associates degree | 0.85 (0.58, 1.26) | 0.422 | 0.78 (0.51, 1.19) | 0.246 |
| Some college | 0.83 (0.58, 1.19) | 0.305 | 0.78 (0.53, 1.14) | 0.195 |
| Bachelor's degree | 1.39 (1.00, 1.92) | 0.049 | 0.96 (0.66, 1.39) | 0.834 |
| Graduate degree | 1.94 (1.35, 2.79) | <0.001 | 1.34 (0.87, 2.06) | 0.188 |
| **Census region** |  |  |  |  |
| Midwest | *Ref.* | . | *Ref.* | . |
| Northeast | 1.52 (1.06, 2.17) | 0.022 | 1.35 (0.92, 1.99) | 0.129 |
| South | 1.21 (0.89, 1.65) | 0.228 | 1.10 (0.79, 1.54) | 0.564 |
| Southwest | 1.36 (0.97, 1.89) | 0.072 | 1.11 (0.77, 1.61) | 0.566 |
| **Political Party** |  |  |  |  |
| Republican | *Ref.* | . | *Ref.* | . |
| Democrat | 0.90 (0.69, 1.18) | 0.456 | 1.15 (0.85, 1.55) | 0.361 |
| Independent | 0.58 (0.43, 0.79) | <0.001 | 0.62 (0.45, 0.86) | 0.004 |
| Something else | 0.25 (0.15, 0.44) | <0.001 | 0.25 (0.13, 0.50) | <0.001 |
| **Household income** |  |  |  |  |
| <\$20,000 | *Ref.* | . | *Ref.* | . |
| \$20,000-39,999 | 1.07 (0.74, 1.54) | 0.739 | 0.96 (0.65, 1.44) | 0.860 |
| \$40,000-\$69,999 | 1.03 (0.73, 1.45) | 0.883 | 0.86 (0.58, 1.25) | 0.406 |
| \$70,000-\$99,999 | 1.69 (1.14, 2.50) | 0.009 | 1.31 (0.83, 2.08) | 0.246 |
| \$100,000-\$149,999 | 2.45 (1.59, 3.78) | >0.001 | 1.82 (1.11, 3.01) | 0.018 |
| > \$150,000 | 2.54 (1.52, 4.25) | <0.001 | 2.08 (1.17, 3.69) | 0.013 |

OR, odds ratio; aOR, adjusted odds ratio

There were numerous reasons provided for being unwilling to receive a COVID-19 vaccine (Table 3). The most common reason was concern about the vaccine’s safety (36.9% 95%CI: 32.8-41.2%), followed by concerns over its efficacy (19.1%, 95%CI: 15.1%, 23.9). There were significant differences in reasons for not being willing to receive the vaccine by age group and political party (See Appendices 4 and 5). There was no significant difference in distribution of reasons by gender, race, education, Census region, and income.

**Table 3.** Reasons for not being willing to receive COVID-19 vaccine.

| **Reason** | **% 95% CI** |
| --- | --- |
| Cost concerns | 4.2 (2.9, 6.0) |
| Concerned vaccine not effective | 17.7 (14.8, 21.1) |
| Concerned vaccine not safe | 38.6 (34.7, 42.7) |
| Concerned about long term health impacts | 15.2 (12.5, 18.4) |
| Vaccine is part of a conspiracy theory | 6.3 (4.6, 8.6) |
| Religious objections | 2.2 (1.3, 3.9) |
| May get it, but don't want to be first* | 2.7 (1.6, 4.5) |
| Do not trust current administration overseeing its development* | 1.4 (0.67, 2.8) |
| Previous side effects or allergies* | 0.8 (0.3, 2.1) |
| Vaccine seen as unnecessary* | 1.6 (0.9, 3.1) |
| Other | 2.0 (1.1, 3.6) |
| Refused | 7.2 (5.4, 9.5) |

\*Recoded from initial response of "Other"

The leading sources of where participants receive vaccine information were their primary care doctor (36.0%), followed by the Centers for Disease Control and Prevention (28.0%), and family (11.2%; See Table 4). There were no significant differences in source of vaccine information by willingness to receive a COVID-19 vaccine. The distribution in sources of vaccination information significantly varied by age, political party, race and census region (Data not shown). There were no differences by gender, education, and income.

**Table 4.**
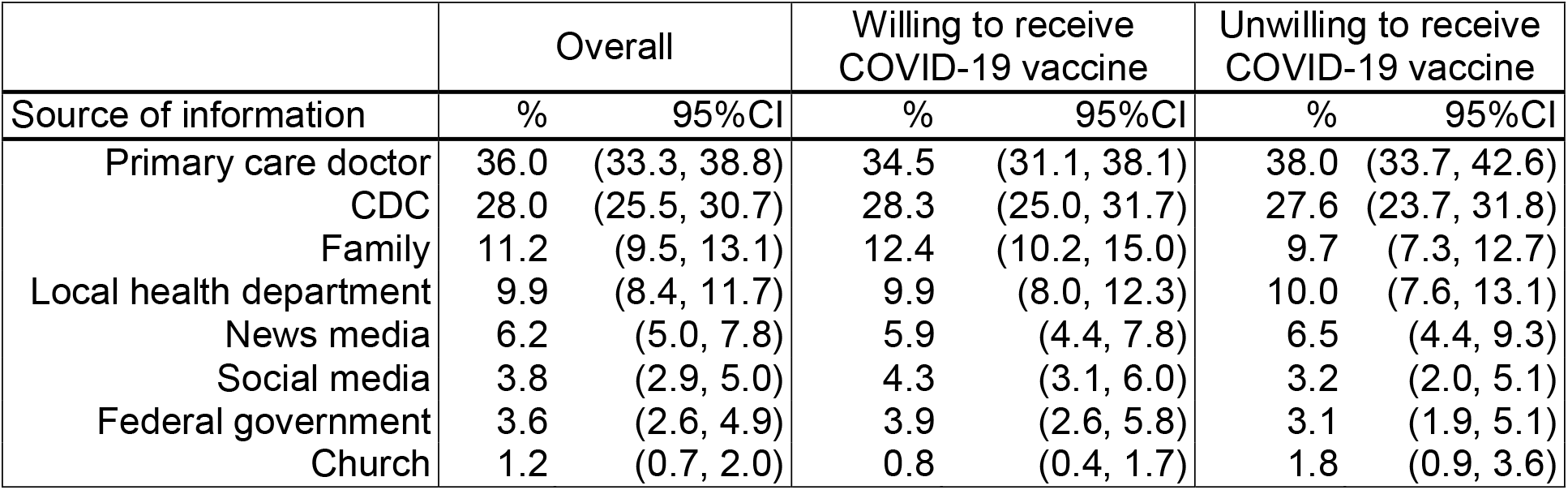
Sources of vaccine information.

## Discussion

Our nationally representative survey results suggest that only 59% of adults across the United States would be willing to receive a COVID-19 vaccine; this would not meet the estimated 70% threshold estimated necessary to achieve herd immunity.^7^ At the time of this survey, a COVID-19 vaccine was not yet available to any segment of the public, other than those enrolled in a clinical trial, and yet 41% of those surveyed expressed an unwillingness to be vaccinated when one was approved. Vaccine willingness differed by race, gender, age, and income level, with African Americans, women, and respondents in the lower income brackets expressing greater reluctance to take the vaccine.

Our study findings reflect a ten percent decrease in the national willingness to accept the COVID-19 vaccine since a study conducted in the early stages of the pandemic (May 2020) through an online survey reported a 69% national willingness to accept the vaccine.^24^ Our results reinforce the results of this prior study which also highlighted lower vaccine acceptance among African Americans, those in the lower income brackets, and those with a more conservative political leaning. This pattern among African American participants is especially concerning, since this segment of the US population continue to bear the highest COVID-19 incidence and mortality rates.^25^ The confluence of chronic disease, other comorbidities and social factors along with COVID-19 has already resulted in disproportionate adverse outcomes among African Americans and Hispanic populations.^19^ High rates of COVID-19 vaccine hesitancy will only further this pernicious disparity, challenging the control of the pandemic among those most affected.

These findings suggest significant obstacles to the successful population-level distribution of candidate COVID-19 vaccines to the US population. To affect substantial reductions in COVID-19 morbidity and mortality, it is important that that the vaccine is swiftly made available to the general public, especially high-risk groups including the elderly, those with pre-existing chronic conditions and those in service-industry and healthcare professions. Societal obstacles preventing widespread coverage of the vaccine will need to be addressed as part of effective policies and programs aiming to distribute the COVID-19 vaccine in the US.

To understand the measures that can be taken to address COVID-19 vaccine hesitancy and facilitate improved coverage, it is helpful to analyze the stated reasons for vaccine unwillingness. Concerns about vaccine safety and efficacy were some of the primarily cited reasons, likely resulting from the uncharacteristically rapid development and testing of the vaccine. These concerns combine with underlying suspicions about vaccination and population-level health interventions led by government among historically marginalized communities of color who have borne the brunt of unethical experimentation, systematic discrimination and systemic, institutional racism. Uniquely, the US pandemic has occurred at a time of historic political and social polarization, characterized by social-media fueled mis- and disinformation. This has driven a further erosion of trust in public institutions, validated by a uncoordinated and heterogenous implementation of federal and state government’s pandemic response. COVID-19 messaging from ‘trusted sources’, including leading politicians and civic leaders have often run contrary to the opinions of leading scientists. Regardless of the causes of the mistrust in COVID-19 vaccinations, this ‘trust’ gap will have to be addressed by federal, state and other health authorities around the country to ensure that the population understands clearly how the unusually expedited discovery, testing and approval process took pains to not circumvent essential measures to ensure vaccine safety, efficacy and risk monitoring.

Vaccine education must be a seminal component of the COVID-19 immunization strategy in the US, to maximize the likelihood of success. A crude, one-size-fits-all approach is unlikely to be effective and risks being counter-productive given the complexity of immunization decision-making and wide variability in vaccine attitudes and beliefs.^26^ A more nuanced approach might be that of message tailoring, which involves providing customized vaccine messages based on an individual’s unique beliefs, experiences, knowledge, and barriers to action.^27^ Research on using a tailoring approach has shown that by increasing the personal relevance for a specific target audience, people are more receptive to new information that may challenge their beliefs. As such, message tailoring can maximize vaccine acceptance.^28^

Our study presents a nationally representative sample, powered to detect differences at the Census region level, as well as among certain racial groups. There are several limitations that are worth highlighting. First, our respondents were identified using a web-based platform, which required access to a smartphone or computer with internet access to participate in the survey. In addition, while our analysis was weighted to capture differences in vaccine acceptance by White, African American, or Hispanic racial groups, we were unable to capture differences among smaller racial/ethical subgroups. Third, we cannot assume that reported willingness to take COVID-19 vaccine (or lack thereof) would result in actual action. Since data collection was conducted in September 2020, before the public release of safety and efficacy results from Phase III clinical trials for several vaccine candidates, it may be possible that our estimates of vaccine willingness may be underestimated given the high efficacy and safety profiles of leading vaccine candidates.^5,6^

In conclusion, we found that a substantial proportion (41%) of United States residents are unwilling to receive a COVID-19 vaccine as soon as one is made publicly available. However, we found that vaccine acceptance differs by sub-populations; with females, African Americans, and older age respondents reporting being less willing to receive the vaccine. This is particularly concerning from a public health perspective given that several of these sub-groups are at highest risk for severe disease outcomes. In addition to sub-group differences in willingness to receive the vaccine, respondents provided a variety of reasons for being unwilling to receive the vaccine, driven by various sources of vaccine information (and misinformation). This compounds the challenge of delivering a safe and efficacious COVID-19 vaccine at a population level to achieve herd immunity. A multi-pronged and targeted communications and outreach effort is likely needed to achieve a high level of immunization coverage. The promise of highly efficacious COVID-19 vaccines to curtail the pandemic in the US, spurring a return to normalcy and economic revitalization will fall short if adequate attention is not paid to these concerning levels of hesitancy to accept the vaccine.

## Data Availability

Abridged datasets may be made available upon written request

## Declaration of interests

We declare no competing interests.

## Data sharing

The data analyzed during the current study are available from the corresponding author on reasonable request.

## Contributors

DGG, SA, and ABL designed the study and developed the study protocol. ABL provided scientific and study oversight. All authors generated hypotheses, interpreted the data, and critically reviewed the manuscript. DGG wrote the first draft of the manuscript and conducted analyses with AM.

## Acknowledgements

We thank colleagues Gregory Kirk, Shruti Mehta, and Sunil Solomon for input on the study design.

## Notes

### Competing Interest Statement

The authors have declared no competing interest.

### Author Declarations

The study protocol and survey instruments were approved by the Institutional Review Board at Johns Hopkins Bloomberg School of Public Health (IRB00012413). All participants provided consent (electronically).

