## Appendices 1-5 for "COVID-19 Vaccine Acceptability and Inequity in the United States: Results from a nationally representative survey"

**APPENDIX 1 Demographic characteristics for Whites, Blacks, and Hispanics**

| **Age category** | **White** | **Black** | **Hispanic** |
| --- | --- | --- | --- |
| 18-24 | 6.9 (5.2, 9.1) | 19.4 (15.7, 23.8) | 14.1 (10.8, 18.3) |
| 25-34 | 20.2 (17.3, 23.4) | 25.6 (21.4, 30.2) | 23.4 (19.3, 28.2) |
| 35-44 | 22.8 (19.8, 26.1) | 15.6 (12.3, 19.5) | 15.5 (12.2, 19.6) |
| 45-54 | 19.4 (16.5, 22.5) | 10.2 (7.7, 13.4) | 12.0 (9.1, 15.7) |
| 55-64 | 13.8 (11.4, 16.5) | 13.3 (10.3, 17.1) | 18.8 (15.0, 23.3) |
| 65+ | 17.0 (14.4, 19.9) | 15.9 (12.5, 20.0) | 16.1 (12.5, 20.4) |
| **Gender** |  |  |  |
| Female | 48.6 (44.8, 52.4) | 54.0 (48.9, 58.9) | 48.3 (43.1, 53.6) |
| Male | 51.1 (47.3, 54.9) | 45.4 (40.4, 50.4) | 51.4 (46.1, 56.6) |
| Other | 0.3 (0.1, 1.2) | 0.7 (0.2, 2.1) | 0.3, (0.1, 2.0) |
| **Educational Attainment,** |  |  |  |
| High school degree or less | 19.1 (16.3, 22.3) | 30.9 (26.4, 35.8) | 21.7 (17.7, 26.3) |
| Associate degree | 13.4 (11.1, 16.2) | 13.1 (10.1, 16.8) | 13.3 (10.2, 17.3) |
| Some college (no degree) | 17.3 (14.6, 20.4) | 18.4 (14.9, 22.6) | 21.6 (17.5, 26.3) |
| Bachelor’s degree | 28.0 (24.8, 31.5) | 23.2 (19.3, 27.7) | 29.9 (25.4, 35.0) |
| Graduate degree | 22.1 (19.1, 25.4) | 13.7 (10.6, 17.6) | 13.1 (10.0, 17.0) |
| **Annual Household Income** |  |  |  |
| < $20,000 | 15.0 (12.5, 17.9) | 24.2 (20.1, 28.9) | 14.3 (11.0, 18.3) |
| $20,000 – $39,000 | 17.1 (14.5, 20.2) | 23.2 (19.2, 27.8) | 22.4 (18.3, 27.1) |
| $40,000 – $69,000 | 27.8 (24.5, 31.3) | 25.2 (21.0, 29.8) | 26.0 (21.6, 30.9) |
| $70,000 – $99,000 | 16.9 (14.3, 19.9) | 11.1 (8.0, 14.6) | 18.1 (14.3, 22.5) |
| $100,000 – $149,000 | 13.8 (11.4, 16.7) | 8.3 (5.9, 11.4) | 14.3 (11.0, 18.5) |
| >$150,000 | 9.3 (7.3, 11.7) | 8.0 (5.7, 11.2) | 4.9 (3.1, 7.8) |
| **Political party** |  |  |  |
| Democrat | 30.7 (27.4, 34.3) | 64.4 (59.4, 69.0) | 47.4 (42.2, 52.7) |
| Republican | 38.7 (35.1, 42.4) | 12.7 (9.7, 16.4) | 24.1 (19.8, 28.9) |
| Independent | 25.8 (22.6, 29.3) | 15.8 (12.5, 19.7) | 22.5 (18.4, 27.3) |
| Something else/Refused | 4.8 (3.4, 6.7) | 7.2 (5.0, 10.3) | 6.0 (3.9, 8.9) |

Caption: Data are weighted % (95%CI)

**Appendix 2 Willingness to receive COVID-19 vaccine when publicly available by subgroup**

|  | **%** | **95%CI** |
| --- | --- | --- |
| **Gender** |  |  |
| Male | 66.1% | (62.4, 69.7) |
| Female | 51.5% | (47.6, 55.4) |
| **Age group** |  |  |
| 18-24 | 64.4% | (56.2, 71.9) |
| 25-34 | 62.7% | (57.0, 68.1) |
| 35-44 | 60.5% | (54.1, 66.6) |
| 45-54 | 52.4% | (45.4, 59.2) |
| 55-64 | 50.4% | (43.4, 57.5) |
| 65+ | 62.6% | (56.1, 68.6) |
| **Race** |  |  |
| White | 59.5% | (55.6, 63.2) |
| Black | 48.8% | (43.7, 54.0) |
| Latino | 56.8% | (51.5, 62.1) |
| Asian | 82.3% | (67.8, 91.1) |
| **Education** |  |  |
| ≤ High school | 54.9% | (48.7, 60.8) |
| Associate's | 50.9% | (43.4, 58.4) |
| Some college | 50.2% | (43.8, 56.6) |
| Bachelors | 62.8% | (57.8, 67.5) |
| Graduate | 70.3% | (64.2, 75.7) |
| **Census region** |  |  |
| Northeast | 63.7% | (57.5, 69.4) |
| Midwest | 53.6% | (47.5, 59.7) |
| South | 58.3% | (53.7, 62.8) |
| West | 61.1% | (55.7, 66.2) |
| **Income** |  |  |
| <20K | 51.8% | (44.9, 58.6) |
| 20K to <40K | 53.3% | (47.2, 59.4) |
| 40K to <70K | 52.4% | (47.1, 57.7) |
| 70K to <100K | 64.4% | (57.8, 70.6) |
| 100K to <150K | 72.4% | (65.3, 78.6) |
| ≥ 150K | 73.2% | (63.9, 80.8) |
| **Political Party** |  |  |
| Democrat | 62.4% | (58.4, 66.3) |
| Republican | 64.8% | (59.8, 69.5) |
| Independent | 51.7% | (46.1, 57.2) |
| Something else | 31.8% | (21.7, 44.0) |

**Appendix 3 ---Comparison of willingness to receive COVID-19 and influenza vaccine**

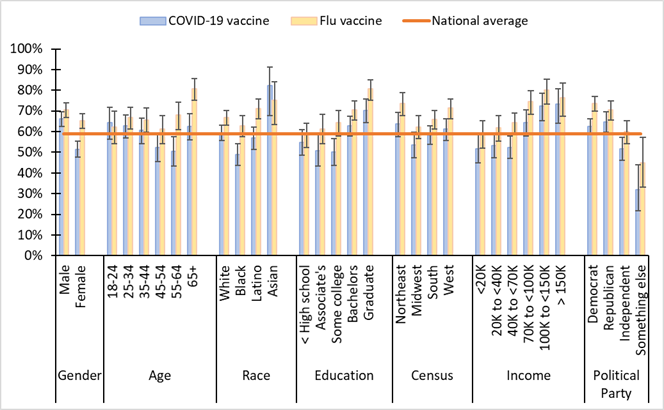

Caption: Data are weighted % (95%CI)

Appendix 4 Reasons for not receiving COVID-19 vaccine by age group

| **Reason** | **18-24** | **25-34** | **35-44** | **45-54** | **55-64** | **65+** |
| --- | --- | --- | --- | --- | --- | --- |
| Cost concerns | 11.4 (5.6, 22.1) | 5.3 (2.7, 10.1) | 6.9 (3.5, 13.0) | 1.5 (0.3, 6.4) | 1.2 (0.3, 6.4) | 1.6 (0.3, 7.4) |
| Concerned vaccine not effective | 22.0 (13.2, 34.3) | 18.6 (12.5, 26.7) | 17.0 (11.4, 24.5) | 17.2 (11.1, 25.8) | 15.6 (9.8, 23.9) | 18.2 (11.3, 27.9) |
| Concerned vaccine not safe | 33.3 (22.8, 45.7) | 36.1 (27.9, 45.2) | 38.9 (30.0, 48.7) | 38.5 (29.6, 48.4) | 43.9 (34.8, 53.4) | 38.7 (29.3, 49.0) |
| Concerned about long term health impacts | 3.9 (1.2, 11.8) | 15.5 (9.9, 23.4) | 11.3 (6.5, 18.8) | 18.5 (12, 27.5) | 23.2 (16.2, 32.0) | 13.7 (8.1, 22.3) |
| Vaccine is part of a conspiracy theory | 10.3 (4.5, 21.8) | 7.7 (4.1, 13.8) | 6.3 (3.1, 12.4) | 10.2 (5.5, 17.9) | 0.5 (0.1, 3.5) | 4.1 (1.7, 9.4) |
| Religious objections | 5.9 (1.9, 16.8) | 3.0 (1.2, 7.5) | 2.9 (1.0, 8.3) | 2.4 (0.7, 7.5) | --- | --- |
| May get it, but don’t want to be first* | --- | 2.2 (0.5, 8.3) | 2.7 (0.8, 8.5) | 2.4 (0.7, 8.2) | 2.9 (1.1, 7.4) | 5.4 (2.0, 13.5) |
| Don’t trust current administration overseeing its development* | --- | --- | 2.1 (0.5, 8.3) | 1.3 (0.2, 8.9) | 0.6 (0.1, 2.7) | 4 (1.4, 10.5) |
| Previous side effects or allergies* | --- | 1.2 (0.2, 8.2) | --- | 0.3 (0, 2.5) | 0.9 (0.1, 5.9) | 2.4 (0.6, 9.2) |
| Vaccine seen as unnecessary* | --- | 0.3 (0.0, 2.2) | 0.9 (0.1, 6.4) | 2.1 (0.5, 7.9) | 3.8 (1.3, 10.4) | 2.3 (0.6, 7.6) |
| Other | 2.6 (0.6, 11.4) | --- | --- | 2.9 (0.9, 9.2) | 2.1 (0.5, 8.0) | 5.7 (2.4, 13.1) |
| Refused | 10.6 (5.3, 19.9) | 10.2 (5.8, 17.1) | 10.9 (6.4, 17.9) | 2.7 (0.9, 7.9) | 5.3 (2.5, 10.8) | 3.9 (1.5, 10.0) |

Caption: Data are weighted % (95%CI)

Appendix 5 Reasons for not receiving COVID-19 vaccine by political party

| **Reason** | **Republican** | **Democrat** | **Independent** | **Something Else** |
| --- | --- | --- | --- | --- |
| Cost concerns | 3.4 (1.6, 7.0) | 4.3 (2.3, 7.9) | 4 (2.0, 7.6) | 7.3 2.7, 18.2) |
| Concerned vaccine not effective | 11.8 (7.5, 18.3) | 21.3 (16.4, 27.1) | 18.1 (12.8, 25.1) | 20.7 (12.0, 33.2) |
| Concerned vaccine not safe | 35.1 (27.8, 43.3) | 41.7 (35.6, 48.1) | 40.7 (33.2, 48.6) | 30.8 (19.1, 45.7) |
| Concerned about long term health impacts | 15.8 (10.6, 22.9) | 14.6 (10.7, 19.5) | 14.6 (9.8, 21.1) | 17.7 (9.4, 30.9) |
| Vaccine is part of a conspiracy theory | 11.5 (7.3, 17.6) | 3.9 (2.3, 6.5) | 5.5 (2.7, 10.7) | 2.1 (0.3, 13.3) |
| Religious objections | 3.2 (1.3, 7.8) | 2.1 (0.9, 4.8) | 1.9 (0.7, 5.4) | --- |
| May get it, but don’t want to be first* | 3.4 (1.4, 7.8) | 1.0 (0.4, 2.6) | 3 (1.1, 7.7) | 6.8 (2.2, 19.1) |
| Don’t trust current administration overseeing its development* | 3.0 (1.4, 7.8) | 1.1 (0.2, 5.1) | --- | 3.0 (1.4, 7.8) |
| Previous side effects or allergies* | --- | 2 (0.7, 5.6) | --- | --- |
| Vaccine seen as unnecessary* | 1.0 (0.3, 3.3) | 1.3 (0.3, 5.1) | 2.1 (0.3, 13.3) | 1.0 (0.3, 3.3) |
| Other | 2.0 (0.7, 5.5) | 2.2 (0.7, 6.1) | --- | 2.0 (0.7, 5.5) |
| Refused | 5.2 (3.0, 8.6) | 5.7 (3.0, 10.4) | 12.7 (6.5, 23.1) | 5.2 (3.0, 8.6) |

Caption: Data are weighted % (95%CI)
